# Dose-Response in Modulating Brain Function with Transcranial Direct Current Stimulation: From Local to Network Levels

**DOI:** 10.1101/2022.11.08.22282088

**Authors:** Ghazaleh Soleimani, Rayus Kupliki, Martin Paulus, Hamed Ekhtiari

## Abstract

**Background:** Non-invasive brain stimulation methods for modulating brain activity via transcranial technologies like transcranial direct current stimulation (tDCS) are increasingly prevalent to investigate the relationship between modulated brain regions and stimulation outcomes. However, the inter-individual variability of tDCS has made it challenging to detect intervention effects at the group level. Collecting multiple modalities of magnetic resonance imaging data (i.e., structural and functional MRI) helps to investigate how dose-response ultimately shapes brain function in response to tDCS.

**Method:** We collected data in a randomized, triple-blind, sham-controlled trial with two parallel arms. Sixty participants with MUD were randomly assigned to sham or active tDCS (n=30 per group, 2 mA, 20 minutes, anode/cathode over F4/Fp1). Structural and functional MRI (including high-resolution T1 and T2-weighted MRI, resting-state fMRI, and methamphetamine cue-reactivity task with meth versus neutral cues) were collected immediately before and after tDCS. T1 and T2-weighted MRI data were used to generate head models for each individual to simulate electric fields. Associations between electric fields (dose) and changes in brain function (response) were investigated at four different levels: (1) voxel level, (2) regional level (atlas-based parcellation), (3) cluster level (active clusters in the contrast of interest), and (4) network level (both task-based and resting-state networks).

**Result:** At the (1) voxel-level, (2) regional level, and (3) cluster level, our results showed no significant correlation between changes in the functional activity and electric fields. However, (4) at the network level, a significant negative correlation was found between the electric field and ReHo in the default mode network (r=-0.46 (medium effect size), p corrected=0.018). For the network-level analysis of task-based fMRI data, frontoparietal connectivity showed a positive significant correlation with the electric field in the frontal stimulation site (r=0.41 (medium effect size), p corrected=0.03).

**Conclusion:** The proposed pipeline provides a methodological framework to analyze tDCS effects in terms of dose-response relationships at four different levels to directly link the electric field (dose) variability to the variability of the neural response to tDCS. The results suggest that network-based analysis might be a better approach to provide novel insights into the dependency of the neuromodulatory effects of tDCS on the brain’s regional current dose in each individual. Dose-response integration can be informative for dose optimization/customization or predictive/treatment-response biomarker extraction in future brain stimulation studies.

## Introduction

Non-invasive brain stimulation methods for modulating brain activity via transcranial technologies are increasingly prevalent to investigate the relationship between modulated brain regions and stimulation outcomes [1]. As one of the most frequently used technologies, transcranial direct current stimulation (tDCS) has shown promising results to modulate brain activity/connectivity in both healthy people and those with neurological/psychiatric disorders [2-4]. Most of the previous electrophysiological and neuroimaging studies showed encouraging results that the injected current by tDCS can change cortical excitability ands brain functions [5, 6].

By considering that brain regions do not operate in isolation but are functionally connected, more researchers have started to investigate the effect of injected current on brain functions in targeted and non-target brain areas [7, 8]. However, the inter-individual variability of tDCS has made it challenging to detect intervention effects at the group level [9] and the main challenge is that potential effect of tDCS is limited by small effect sizes [10-12]. Different sources of variability including anatomical (e.g., fat thickness, skull thickness, amount of CSF that affect current flow through the brain) and functional factors (e.g., inherent oscillations, ongoing brain activity or connectivity that affect brain responses to the injected current) have been identified for tDCS studies [9, 13-15]. On one hand, advances in neuroimaging techniques provide new insights into the effects of functional state in response to tDCS [16]. On the other hand, as it is impossible to non-invasively measure the strength of tDCS-induced electric field (EFs), the EFs are often modeled computationally [17] which are validated repeatedly by intracranial recording [18, 19], physiological [20], and neuroimaging [21, 22] studies. Computational head models showed that delivered dose into the cortical (amount of current reaching the brain) is different from the delivered dose into the scalp (e.g., fixed 2 mA current intensity) and varies across the population based on individualized haad and brain anatomy. Under the assumption that EF intensity over the cortex relates to the tDCS responses at the functional level, the association between stimulation dose and brain responses at a cortical target site could explain dose-response relationships in tDCS studies.

Collecting multiple modalities of magnetic resonance imaging data (i.e., structural and functional MRI) helps to investigate dose-response relationship and define how tDCS induced EFs may change underlying brain functions. To this end, high resolution structural MRI are commonly used for creating computational head models (CHMs) to predict EF distribution patterns over the cortex. Meanwhile, fMRI data aims to capture the cortical functional activity in response to the injected current. However, there have been few studies dedicated to investigating the dose-response relationships based on integrating CHMs with fMRI data.

One of the earliest studies in which both EFs and fMRI data were discussed was in Halko et al 2011. In that study, a single-subject case study of tDCS with combined visual rehabilitation training after stroke revealed that EF intensity obtained from CHMs correlated with task-based fMRI activation in some predefined ROIs based on using voxel-wise correlation analysis within the brain regions between two maps. In another study, CHMs were generated for a group of participants with left-sided glioma, and the averaged EF strength was extracted from the left and right M1 ROIs, and a significant correlation between averaged EFs in the right M1 and changes in global connectivity from the right M1 was reported [23]. A similar correlational approach was also used by Antonenko et al. to assess the relationship between tES-induced EFs and neurophysiological outcomes [24]. In that study, three 4 mm spheres were defined around the left precentral gyrus. Two different EF components, including tangential and normal components, were extracted from each ROI and analyzed separately to test for correlations with tDCS-induced functional coupling in sensorimotor network and significant negative/positive correlations were reported between tangential/normal component of the EF and resting-state functional connectivity [24]. Esmielpour et al used atlas-based parcellation for defining ROIs over the CHMs and fMRI data during a drug cue reactivity task in a group of participants with methamphetamine use disorder and reported a significant correlation between changes in brain activation and averaged EF intensity only in frontal pole as the area that received maximum EF compared to other predefined regions [25]. In another study conducted by Jamil et al, in order to assess whether cerebral blood flow (CBF) activations across the cortex obtained from fMRI agree respectively with EFs obtained from CHMs, a voxel-wise rank correlation was used to calculate the relationship between averaged EF distribution patterns in MNI space and the group-level T-contrast images for each active tDCS intensity obtained from a whole-brain analysis and significant correlations between EFs and changes in CBF were reported at the voxel-level in both cathodal and anodal stimulation [26]. Recently it has also been shown that current density in the left DLPFC positively correlated with changes in functional connectivity between two predefined ROIs (left dorsolateral and left ventrolateral PFC) during a working memory task based on using psychophysiological interaction analysis and simulating precise computational head models.

Previous dose-response relationship papers are limited into two major regional levels to examine the associations between EFs and brain functions; (1) voxel-based, and (2) ROI-based. However, there are other levels of brain response to explore the relationships between dose (EFs) and response (brain functions as measured with fMRI) within brain circuits or networks. Therefore, in this study, we present our pre-registered trial data (NCT03382379) on the association between individualized dose (electric fields estimated with subject-specific computational head models) and brain response (changes in BOLD signals) in a group of participants with methamphetamine use disorders (MUDs) in multiple level. In an exploratory approach, here, we propose that the dose-response relationship could be investigated in at least 4 different levels. Including (1) voxel-level, (2) region-level, (3) cluster-level, and (4) network-level. Considering highly connectedness nature of brain, we anticipated associations between EFs and functional changes in the brain would be stronger at the network-level compared to other levels of associations.

## Method

### Participants

Participants included 60 participants (all-male, mean age ± standard deviation (SD) = 35.86 ± 8.47 years ranges from 20 to 55) with methamphetamine use disorder (MUD). All participants were recruited during their early abstinence from the 12&12 residential drug addiction treatment center in Tulsa, Oklahoma in the process of a clinical trial to measure the efficacy of tDCS on methamphetamine craving (ClinicalTrials.gov Identifier: NCT03382379). Written informed consent was obtained from all participants before the scans and the study was approved by the Western IRB (WIRB Protocol #20171742). This study was conducted in accordance with the Declaration of Helsinki and all methods were carried out following relevant guidelines and regulations. More details on inclusion and exclusion criteria can be found here [27].

### Data collection procedure

Data were collected in a randomized, triple-blind, sham-controlled trial with two parallel arms. Sixty participants with methamphetamine use disorders (MUDs) were randomly assigned to sham or active tDCS (n=30 per group, 2 mA, 20 minutes, anode/cathode over F4/Fp1). Participants in the active group received 2 mA stimulation during 1140 sec whereas participants in the sham group received 40 sec of 2 mA stimulation. Fade out in the sham group followed by 1100 sec without any stimulation (just impedance was controlled that average current overtime is not more than 2 *μ* A). Structural and functional MRI (including high-resolution T1 and T2-weighted MRI, resting-state fMRI, and methamphetamine cue-reactivity task with meth versus neutral cues) were collected immediately before and after tDCS. T1 and T2-weighted MRI data were used to generate head models for each individual to simulate electric fields (Figure 1). More details on neuroimaging and tDCS parameters can be found here [27].

**Figure 1.**
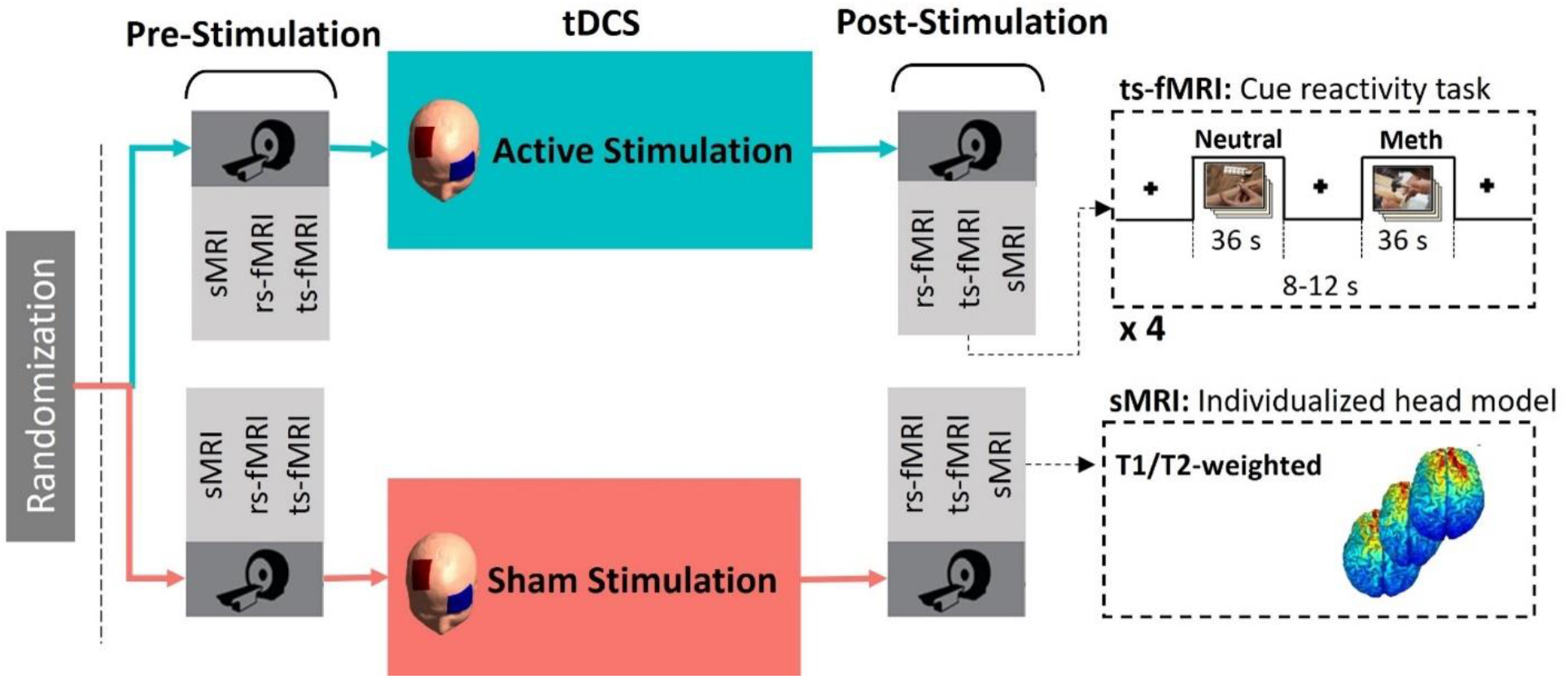
Data acquisition procedure. In a randomized thriple blind sham-controlled clinical trial with a parallel deign, 60 participants with methamphetamine use disorders were randomly assigned to the active or sham groups to receive 20 minutes of unilateral tDCS over DLPFC (anode/cathode over F4/Fp1 in EEG standard system with thwo large 5×7 cm electrode pads). MRI data including structural MRI, resting-state fMRI, and task-based fMRI (a standard cureactivity task) were collected immediately before and after active or sham stimulation. Structural MRI were used for creating individualized computational head models as an indicator of stimulation dose and fMRI data were considered s brain response to the injected curret.

### fMRI data preprocessing

fMRI preprocessing was performed in AFNI. The first three pre-steady state images were removed. The preprocessing steps were despiking, slice timing correction, realignment, transformation to MNI space, and 4 mm of Gaussian FWHM smoothing. Three polynomial terms and the six motion parameters were regressed out. TRs with excessive motion (defined as Euclidian norm of derivative of the six motion parameters being greater than 0.3) were censored during regression.

### Computational head modeling

Gyri-precise CHMs were generated from a combination of high-resolution T1- and T2-weighted MR images for 60 participants. Head models were generated for all participants to visualize how current flows through the brain using SimNIBS software. Briefly, automated tissue segmentation was performed in SPM12. The head volume was assigned to six major head tissues (white matter (WM), gray matter (GM), cerebrospinal fluid (CSF), skull, scalp, and eyeballs). The assigned isotropic conductivity values were WM = 0.126 Siemens/meter (S/m), GM = 0.275 S/m, CSF = 1.654 S/m, skull = 0.01 S/m, skin = 0.465 S/m, and eyeballs = 0.5 S/m. The results were visualized using Gmsh and MATLAB.

### Association analysis

A sample size of 30 participants per arm provided 80% power to detect an effect size (Cohen’s d) of 0.74. Pearson correlation coefficient with FDR correction was used to investigate the associations. As described in the following paragraphs, associations between electric fields (dose) and changes in brain function (response) were investigated at four different levels: (1) voxel level, (2) ROI-level, (3) cluster-level, (4) network-level (both resting-state and task-based).

#### Voxel-level associations

Block design analysis was performed for meth > neutral contrast in pre and post-stimulation scanning sessions based on general linear models (GLM). Functional map and CHMs were transformed to MNI space and non-brain voxels were excluded. EFs were resampled to have identical resolution with functional map and final maps contained about 900k (96 × 114 × 82) voxels. Three-dimensional voxels were vectorized along with the different columns of a matrix for each map where participants were stacked along the rows. Let Xi and Yi be the column vectors across all participants for i ^th^ voxel from EF and fMRI matrices. A whole-brain voxelwise correlation was calculated between each corresponding voxel within the brain. Spatial distribution of the correlations (R-values and p-values) was used for reconstructing the brain map.

#### Region-level associations

For functional activity analysis at the voxel-level, linear mixed-effect model (LME) in AFNI package, which accounts for non-independence of observations inherent in repeated measure data, was used to study the effects of tDCS on the whole brain functional activation. For ROI-level analysis, brain activation during the drug cue reactivity task was calculated for meth > neutral contrast before and after stimulation in each group (real/sham) separately. For each subject mean beta weight values were estimated for all extracted ROIs using Brainnetome atlas (BNA) [28] to show the level of changes (post minus pre-stimulation) task-related activity during meth versus neutral condition and direction of changes was compared between two groups. Change in the ROI-wise activation over time was analyzed with an LME using “nlme” package for linear mixed modeling in R software (v.1.2.5) and included fixed effects of time, group, and interaction between group and time. By-subject intercept and ROI terms (246 ROIs in BNA) were entered as random effects.

In order to determine the relationship between induced EF in each brain region and changes in neural activation (i.e., functional activity after tDCS compared to before stimulation) in response to cue exposure, the correlation between EF strength and changes in functional activity was calculated in both active and sham groups. Statistical results were corrected for multiple comparisons based on false discovery rate correction (FDR).

#### Cluster-level associations

A linear mixed-effect model was used to study the effects of tDCS on the whole brain functional activation during cue exposure. As the fixed effect, time (pre and post-stimulation), group (sham and active), and interaction between them were included in the model. The subject was also considered as a random effect. Group-level functional t-map was calculated in MNI space. Family-wise error (FWE) was found by Monte-Carlo simulation-based (3dClustSim, AFNI) multiple comparison correction with alpha > 0.1. P < 0.005 and cluster size > 40 was considered for reporting the results. Our results revealed five main significant clusters. Averaged BOLD signals were extracted from each cluster. Cluster masks were also applied to the CHMs and averaged EF intensity was extracted from each cluster. Correlations between EF and functional map obtained from the LME model and EFs were calculated at the cluster level.

#### Resting-state network-level associations

CHMs were transformed to MNI space. Regional homogeneity (ReHo) was used for integrating CHMs with resting-state fMRI data at the network level. After preprocessing the functional maps with a routine pipeline in AFNI, the functional images were realigned into the standard MNI space. A bandpass filter (0.01-0.09) was applied and the linear trend was removed. Kendall’s coefficient of concordance (KCC) value was used to calculate the similarity of the time series of a given voxel to its nearest 25 neighbor voxels within a functional cluster (25 nearest neighboring voxels were defined as a cluster and a KCC value was given to the voxel at the center of this cluster). ReHo map was generated for each individual in pre and post-stimulation scans using AFNI software in a voxel-wise fashion. Post minus pre-stimulation map was calculated for each participant in active and sham groups and standardized into z score (Z-ReHo). Each map was combined with the MNI mask to ensure analysis did not include signals from non-brain or white matter voxels. Finally, Z-ReHo maps were smoothed by a Gaussian filter of 4 mm of full width at half maximum (FWHM). A significant threshold was set at corrected P < 0.05 (determined by Monte Carlo simulations with the AFNI AlphaSim program).

Functional atlas parcellation was used for integrating CHMs with Z-ReHo over large-scale brain networks. The Yeo7-2011 atlas was used for extracting the topology of the seven resting-state networks, including visual, somatomotor, dorsal attention, ventral attention, limbic, frontoparietal, and default mode network. Averaged EF intensity and zReHo values were extracted from each large-scale network. Finally, Pearson’s correlation coefficients were computed between the averaged zReHo signal change (post minus pre-stimulation) of large-scale network and EF intensity.

#### Task-based network-level associations

In order to perform seed-to whole-brain task-based functional connectivity, seed regions were defined based on maximum EF. CHMs were generated for each individual and transformed to fsaverage space for calculating averaged EF across the population. 99^th^ percentile of the maximum EF was determined over averaged EF map and a 10 mm sphere was defined around its location. This sphere was used for seed-to-whole brain generalized psychophysiological interaction (gPPI) voxels in the brain that alter their connectivity with a seed region of interest in a given context (here the drug cue reactivity task). The averaged BOLD signal was extracted from the seed region and task-modulated functional connectivity was calculated. For first-level analysis, the design matrix included the seed region’s time course, cue exposure task time course, the interaction between task and BOLD signal in the seed. For the second-level analysis time (pre vs. post-stimulation) x group (active vs. sham) interaction was calculated. Voxel-level threshold was P uncorrected < 0.01 and cluster-level threshold was P FDR corrected < 0.05.

## Results

### Voxel-level results

At the whole brain voxel-level, block designed analyses have not found any significant correlation between electric fields and BOLD signal change—post minus pre-stimulation (p corrected>0.05; 9.74% of the voxels with small effect size (0.1<|*r*|<0.3), 1.36% with medium effect size (0.3<|*r*|<0.5), and only 0.09% with large effect size (0.5<|*r*|<1), the total number of voxels were 8530021 and Pearson correlation coefficient (|*r*|) was considered as a measure of effect size). Because of the large number of voxels, FDR correction was applied and none of them survived FDR multiple comparison corrections (Figure 2).

**Figure 2.**
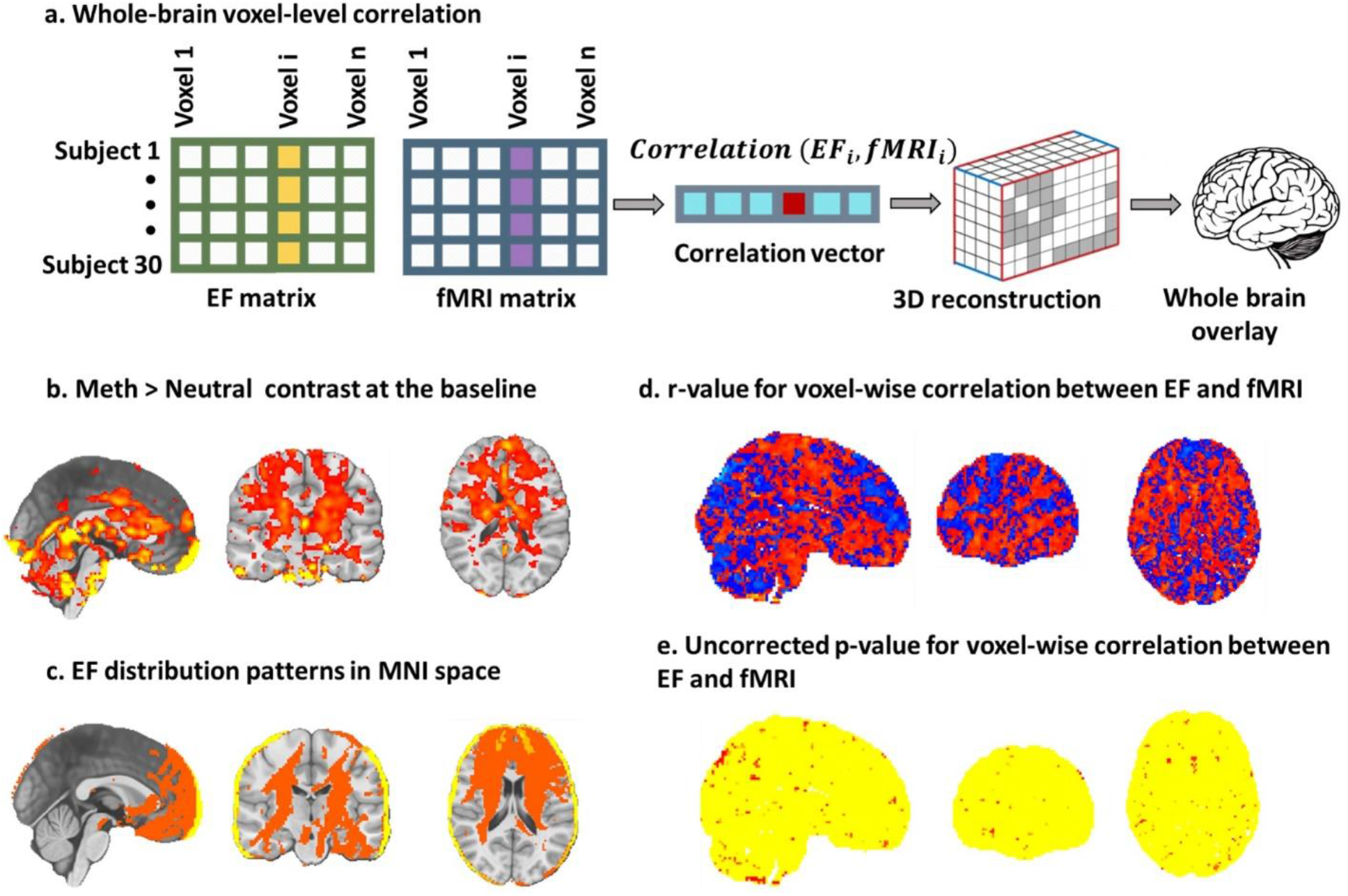
Dose-Response (EF-fMRI correlation) at the voxel-level in a whole-brain approach. **a**. Electric field (EF) maps were vectorized and placed along the columns and participants in the active group along the rows to construct EF matrix. A similar approach was used for generating an fMRI matrix related to the Meth > Neutral contrast. The total number of participants was 30 participants in the active group and the total number of voxels was 897408 voxels. The correlation vector has information about the correlation between each corresponding voxel in EF and fMRI matrices. **b**. Group-level functional map in pre-stimulation is visualized over the MNI space. **c**. EF distribution patterns at the group-level in MNI space. **d**. The voxel-wise correlation coefficient between −1 (cold colors) and 1 (hot colors) over the MNI space. **e**. P values obtained from correlation analysis are mapped over the MNI space. P uncorrected < 0.05 (maximum cluster size < 40) is represented in red. 9.74% of the voxels with small effect size (0.1<|*r*| <0.3), 1.36% of the voxels with medium effect size (0.3< |*r*| <0.5), and only 0.09% of the voxels with large effect size (0.5< |*r*| <1), total number of voxels were 8530021 and *r* was considered as a measure of effect size.

### Region-level results

As shown in Figure 3, at the whole brain regional level, no significant correlation survived FDR correction (24.29% of the regions with small effect size, and 3.33% with medium effect size, the total number of regions were 210 cortical regions).

**Figure 3.**
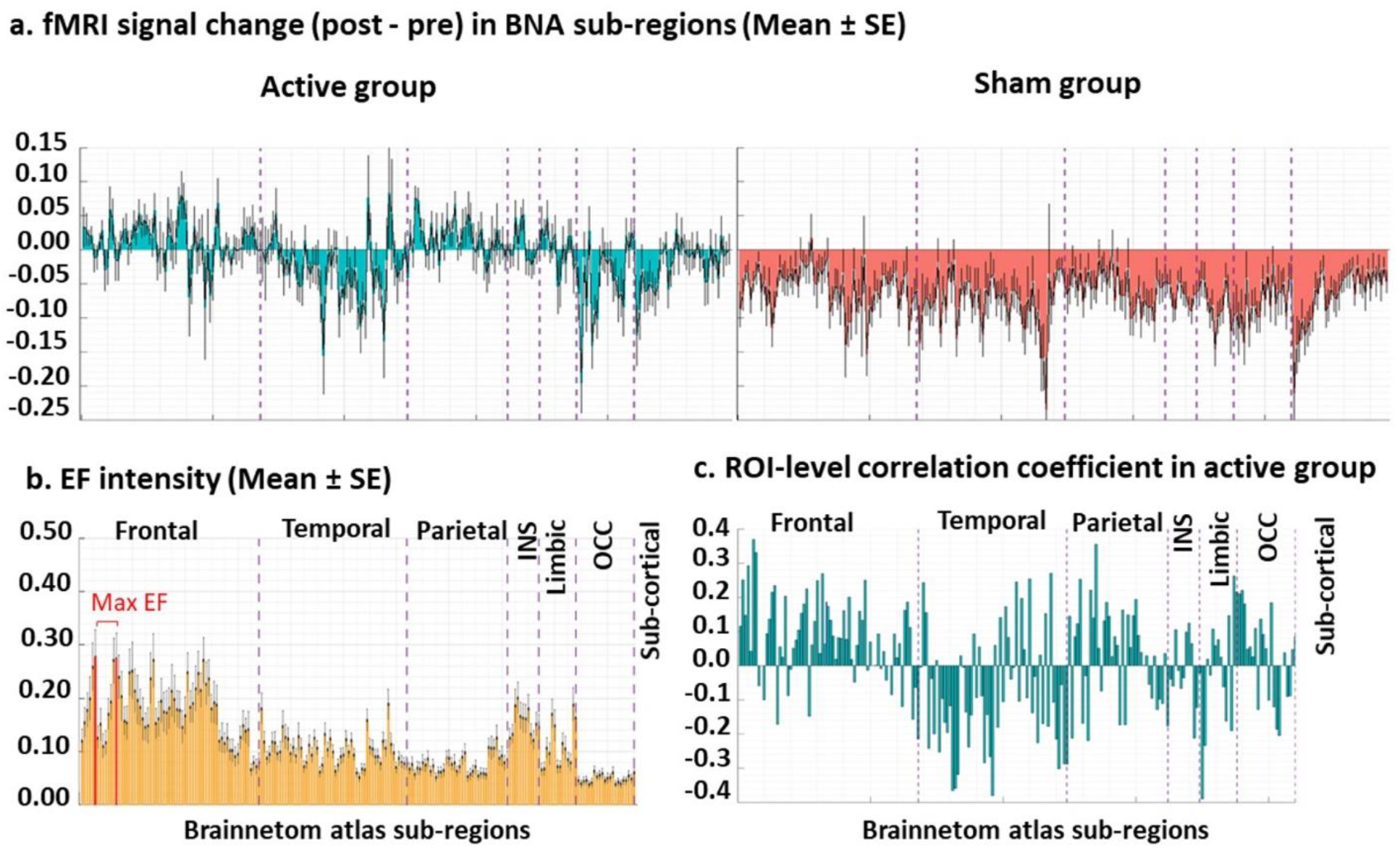
Dose-Response (EF-fMRI correlation) at the Regional-level integration. **a**. First-level functional maps obtained from general linear model (Post minus pre-stimulation) and **b**. individualized computational head models were parcellated using Brainnetome atlas. electric fields (EFs) were calculated only in cortical subregions. **c**. Correlations between all pairs of ROIs were calculated in the active group. After FDR correction, no significant results were achieved. 24.29% of the regions with small effect size, 3.33% of the regions with medium effect size, the total number of regions were 210 cortical regions obtained from Brainnetome atlas parcellation.

### Cluster-level results

At the cluster level (Figure 4), our results showed no significant correlation between changes in the functional activity and electric fields within the clusters (p FDR corrected>0.05; 40% of the clusters with small and 40% with medium effect size; the total number of clusters were 5).

**Figure 4.**
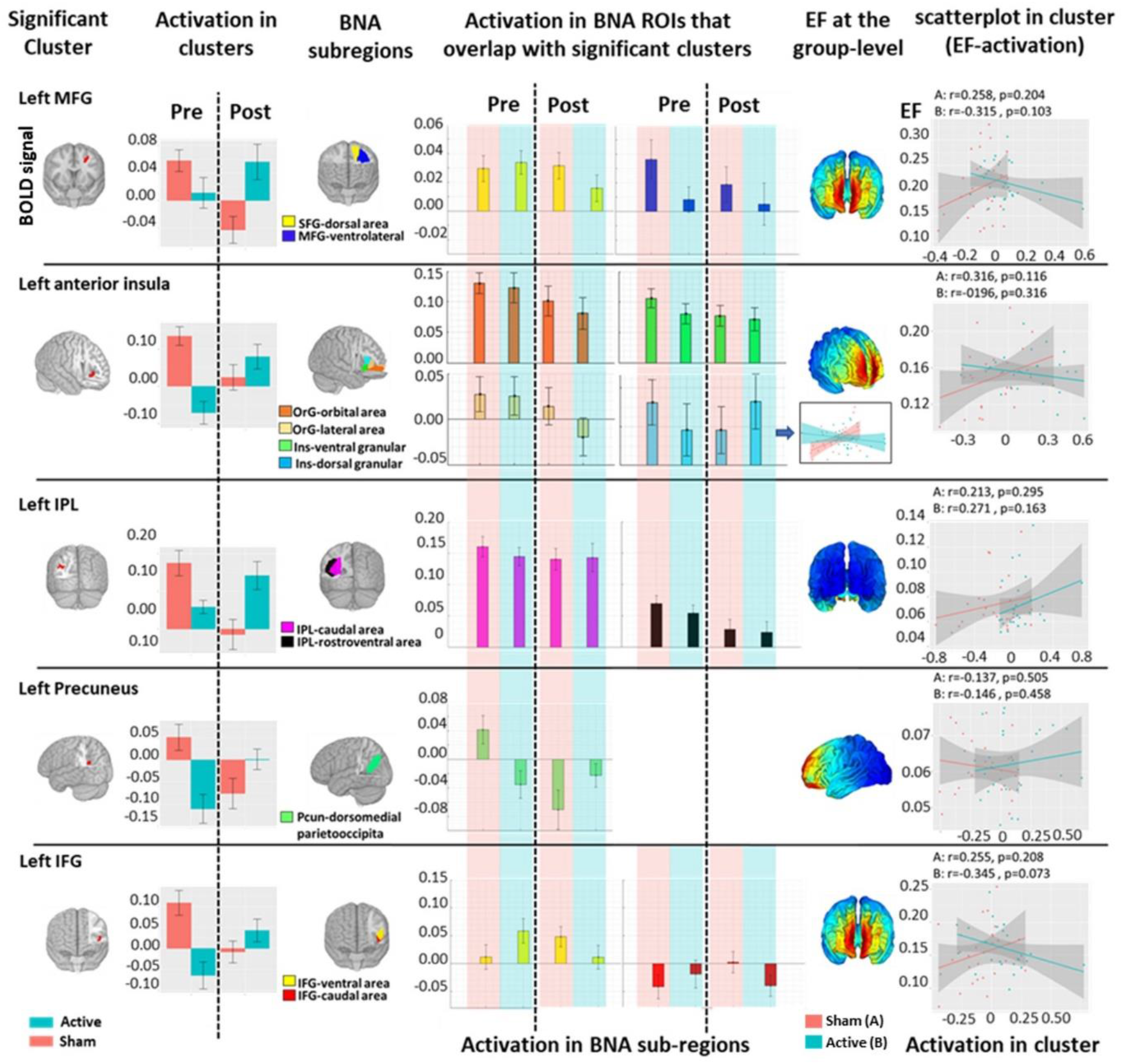
Dose-Response (EF-fMRI correlation) at the Cluster-level integration. <u>First column</u>: Clusters with significant time by group interaction. <u>Second column</u>: BOLD signal within significant clusters before and after stimulation in each group separately. <u>Third column</u>: Brainnetome atlas (BNA) sub-regions that overlap with significant clusters. <u>Fourth column</u>: BOLD signal within BNA sub-regions overlap with significant clusters before and after stimulation in each group separately. <u>Fifth column</u>: Group-level computational head models. <u>Last column</u>: correlation between BOLD signal and EFs within clusters as well as BNA subregions were calculated and cluster level correlations are represented in the last column. No significant correlation was found at the cluster level however in regression analysis significant effect was found for the group. In ROI level correlation significant correlation (without any correction) was found in dorsal angular insula only in the active group (active: r = 0.49 and P uncorrected = 0.0083, sham: r = −0.19 and P uncorrected = 0.33; no significant results after FDR correction). 40% of the clusters with small and 40 of the clusters with medium effect size; the total number of clusters were 5 obtained from time by group interaction in meth > neutral contrast.

### Resting-state network-level results

At the network level (Figure 5), a significant negative correlation was found between electric field and ReHo only in the default mode network in the active group (r=-0.46 (medium effect size), p corrected=0.018). Correlation results were not significant in other extracted large-scale networks.

**Figure 5.**
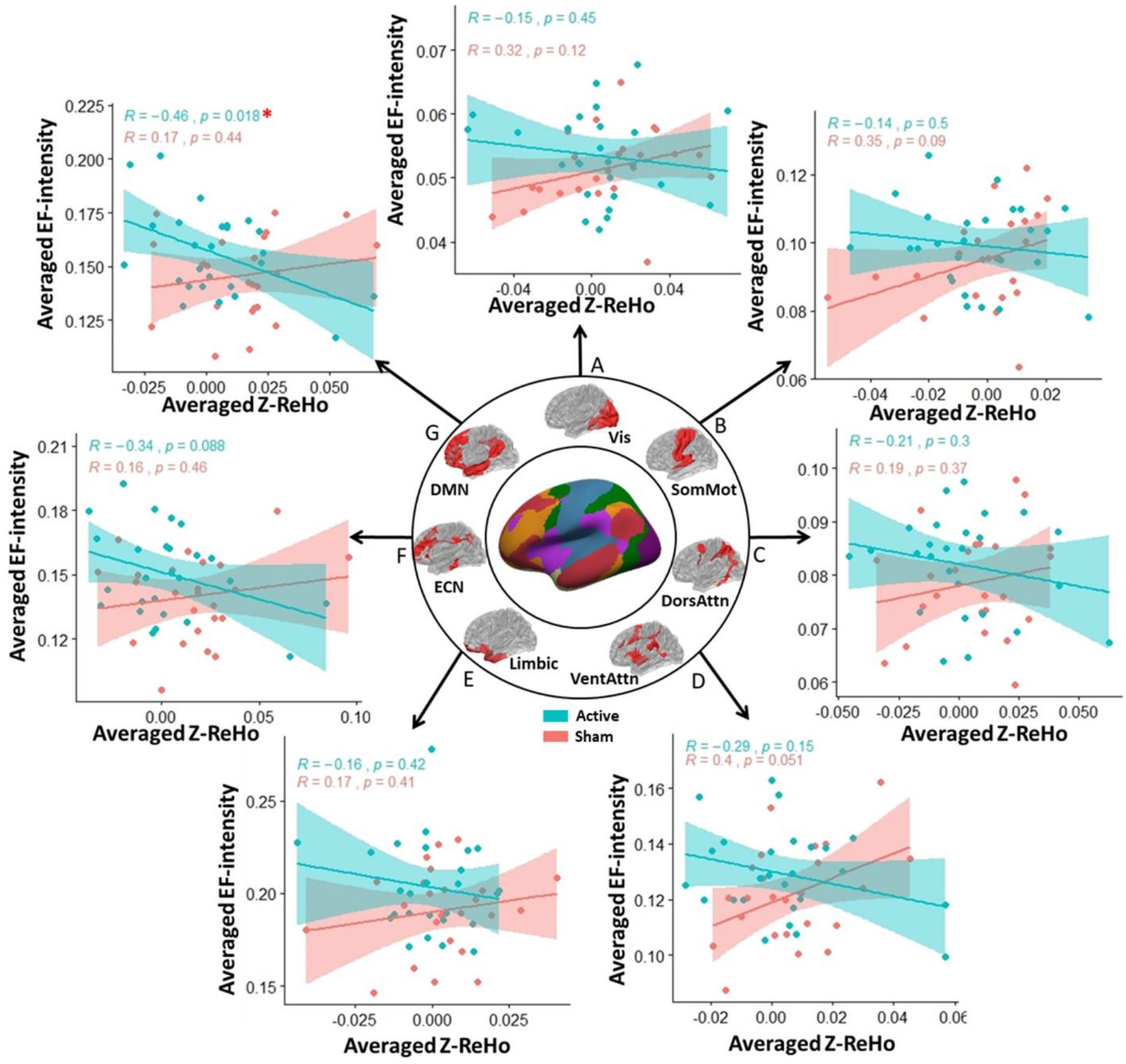
Dose-Response (EF-fMRI correlation) at the Network-level integration. Scatterplots for the correlation between averaged EF intensity and averaged Z-transformed ReHo map within the large scale networks obtained from Yeo7-2011 atlas (A: visual (Vis), B: Somatomotor (SomMot), C: Dorsal attention (DorsAttn), D: Ventral attention (VentAttn), E: Limbic, F: frontoparietal (ECN), G: Default mode (DMN) network). Pearson correlation coefficients and p values for each group are reported above scatterplots. Our results showed significant negative correlation between EF and Z-ReHo maps within the default mode network in the active group (active: r = −0.46 and P FDR corrected = 0.018, sham: r = 0.17 and P FDR corrected = 0.44). Other correlations did not survive FDR correction.

### Task-based network-level results

Our gPPI results (Figure 6) showed two significant clusters as follows: (1) located in bilateral amygdala and hippocamp (cluster size: 1356, (x, y, z) peak coordinate in MNI space: (18,-2,-30) and t-value: 6.59), and (2) located in right inferior parietal lobule (IPL) (cluster size: 701, (x, y, z) peak coordinate in MNI space: (46,-38,38), and t-value: 5.20). ROI-to-ROI gPPI connectivity was also calculated between seed region and each cluster and correlation between averaged EF in seed region and seed-to-cluster task-based connectivity was calculated. For the network-level analysis of task-based fMRI data, frontoparietal connectivity showed a positive significant correlation with electric field in the frontal stimulation site(r=0.41 (medium effect size), p corrected=0.03).

**Figure 6.**
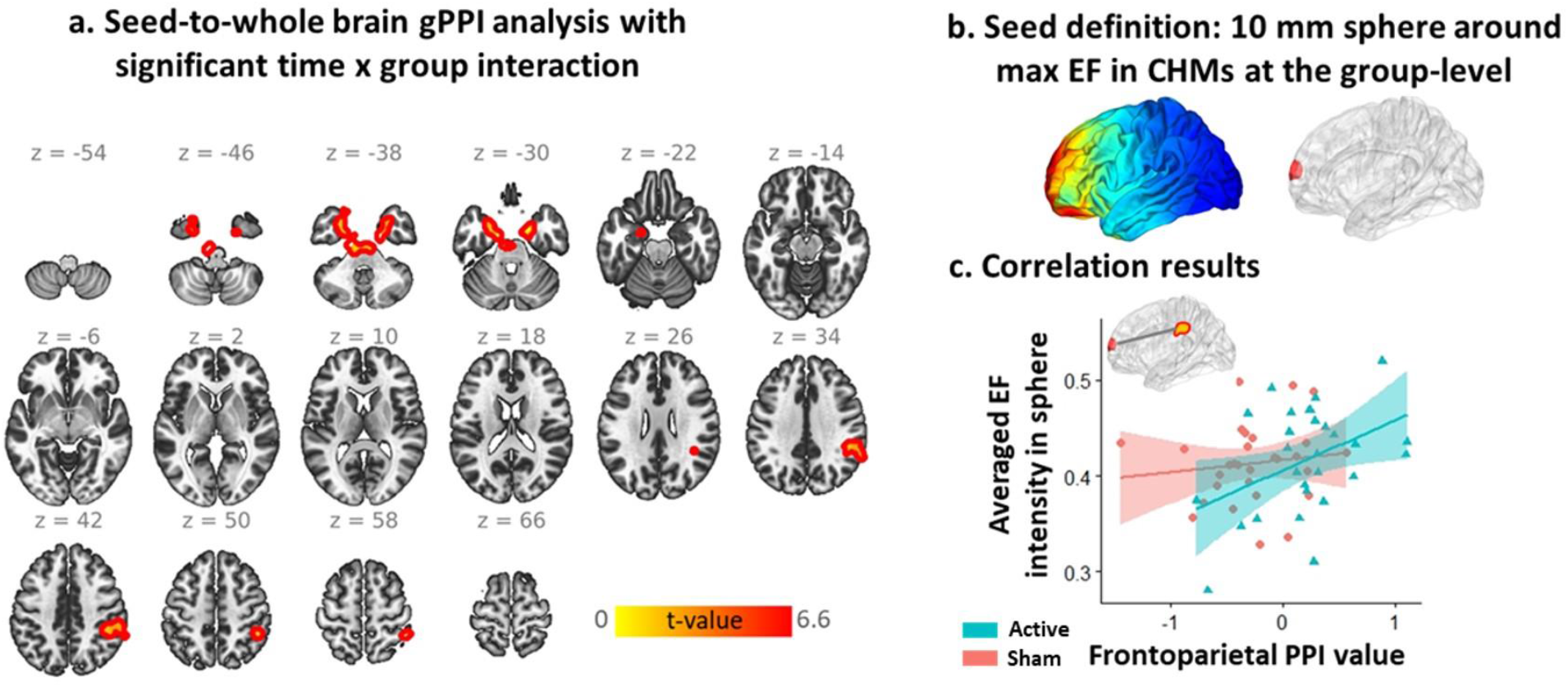
Dose-Response (EF-fMRI correlation) at the Network-level integration based on seed-to-whole brain generalized psychophysiological interaction (gPPI). **a**. Brain regions with significant time by group interaction in gPPI analysis (two significant clusters in bilateral amygdala and hippocamp, and right inferior parietal lobule (IPL)). **b**. defining seed region based on a 10 mm sphere around maximum electric field (EF) location. **c**. correlation between averaged EF intensity within the 10 mm sphere and PPI connectivity between sphere and IPL cluster in each group separately (active: r = 0.41, P corrected = 0.03, sham: r = 0.13, P corrected = 0.52).

## Discussion

Here we investigated the associations between tDCS-induced EFs (dose) and changes in functional activity in response to tDCS based on integrating computational head models and fMRI data. Our results showed that (1) at the whole brain voxel-level, block designed analyses have not found any significant correlation between electric fields and BOLD signal change—post minus pre stimulation (p corrected>0.05; 9.74% of the voxels with small effect size (0.1<|*r*|<0.3), 1.36% with medium effect size (0.3<|*r*|<0.5), and only 0.09% with large effect size (0.5<|*r*|<1), total number of voxels were 8530021 and Pearson correlation coefficient (|*r*|) was considered as a measure of effect size). (2) at the whole brain regional level, no significant correlation survived FDR correction (24.29% of the regions with small effect size, and 3.33% with medium effect size, total number of regions were 210 cortical regions). (3) at the cluster level, our results showed no significant correlation between changes in functional activity and electric fields within the clusters (p FDR corrected>0.05; 40% of the clusters with small and 40% with medium effect size; total number of clusters were 5). (4) at the network level, a significant negative correlation was found between electric field and ReHo in default mode network (r=-0.46 (medium effect size), p corrected=0.018). For the network-level analysis of task-based fMRI data, frontoparietal connectivity showed a positive significant correlation with electric field in the frontal stimulation site(r=0.41 (medium effect size), p corrected=0.03).

### Voxel-level integration

Whole brain voxel-wise correlation across the population showed no specific pattern of associations over the cortex. There is only one previous study that used voxel-wise correlation between EFs and changes in brain functions (using CBF data) and reported a significant correlation between EF distribution patterns in a standard brain and group-level functional map [26]. Despite what we have done here (computed correlation between individualized EF and BOLD signal for each person separately), in that study, one correlation was calculated for all voxels in a single standard head model in MNI space and group-level functional map obtained from group-level analysis of fMRI data. [26].

Dose-reposne relationship at the voxel-level has several challenges. For example, it would be important that two maps be in the same space (subject space or averaged standard space) with the same resolution (based on resampling the maps to have the same resolution as suggested by [29]) to calculate correlation/regression between all brain corresponding voxels in each map across populaiton. Furthermore, integrating results obtained by voxel-wise analysis are inherently limited by the signal-to-noise ratio (SNR) of individual voxel data, which is typically low [30]. Additionally, voxel-wise integration involves a large number of voxels and requires a huge number of testing for any statistical inferencing. Therefore, p-values should be adjusted for multiple comparisons and statististical power would be low at this level of associations.

### Region-level integration

At the whole brain region-level, although our results showed strong correlations in some specific brain regions but no regions survived FDR correction. Almost all of previous dose-response relationship studies were performed within the predefined regions of interest and significant correlations were reported [1, 8, 24, 29]. Different factors can affect associations at this level. For instance, brain regions in CHMs and functional map can be defined based on different approaches, including standard anatomical/functional atlas-based parcellation or placing small spheres around specific brain coordinates (e.g., a 10 mm sphere around maximum EF or specific anatomical region suggested by previous studies according to prior hypothesis and research question). Defining ROIs would be critical to find significant associations between dose and response. For example, Esmaielpur et al reported only one significant correlation which is located in frontopolar cortex that received highest EFs across ten different brain regions. In that study it was shown that parcellation method and defining ROIs is an importan factor for finding significant results in dose-response relationship studies [8].

In dose-response relationship at the region-level features should be extracted from each ROI. In this way, estimated parameters (e.g., beta activation map and EF intensity) are extracted by averaging across the voxels inside the predefined ROIs to calculate the relationships between fMRI-derived parameters and tES-induced EFs. ROI-based integration overcomes the low SNR mentioned in the voxel-wise manner based on averaging data from multiple voxels. In the ROI-based approach, since the statistical testing is performed on ROIs rather than voxels, the total number of tests is reduced. However, results in brain regions other than those already hypothesized and predefined will be lost, and selecting appropriate brain regions would be challenging and needs specific knowledge about the problem for defining relevant areas.

### Cluster-based integration

Our results showed no significant correlation between EFs and changes in functional activity within the significantly modulated areas with tDCS (activated clusters). However, a significant effect of group was found in all five clusters. Although no previous study reported the cluster-based associations, this approach is a method between voxel-based and ROI-based integration such that each voxel has a priority chance of being selected for integration; however, the analysis will be performed in contiguous clusters of voxels similar to the ROI-based approach. In cluster-based integration, fMRI analysis results (e.g., activated cluster in response to a specific task) are used to define ROI. Cluster mask will be applied to CHMs for extracting averaged EF components. This approach is more reliable than atlas-based anatomical landmarks since currently activated brain regions are more vulnerable to the induced EF.

### Network-based integration

With respect to the diffuse current flow in brain stimulation studies, stimulation outcomes can be understood as modulation of global networks. Furthermore, brain regions do not operate in isolation and many brain regions interact with eachother such that brain is seen as a connectom of interacting regions [31]. So, as we hypothesized, we found significant dose-response relationships at the network-level rather than local associations (region-based, voxel-based, or cluster-based approaches). Our results revealed a significant correlation between EF and fMRI data at the large-scale resting-state network-level. We calculated the association between dose and changes in the resting-state networks based on the connectivity within the large scale networks (network atlas based ReHo) and a significant negative correlation were found within the default mode network. No significant results were found within the other networks. The main resons for finding significant associations only in DMN (and not other brain networks) might be related to strong EFs in this network [32]. DMN has a large node in the prefrontal cortex between anode and cathode location where maximum EFs is located.

Since each network covers a widely distributed area, finer sub-regions of the network can be extracted as ROIs, and integration can be performed in the main nodes of the large-scale networks. In this context we also found significant dose-response relationships within the frontoparietal network (which is also named executive control network or ECN). We found a significant positive correlation between task-based frontoparietal functional connectivity and EF intensity within the prefrontal cortex. Our result is in line with previous findings that reported by Indhastrial et. al. during a working memory task [33]. They showed a significant correlation between median and maximum values of the currenct density in DLPFC and changes in task-based functional connectivity in the working memory network in a group of older adults [33].

Two main points should be highlighted based on our results. (1) Our findings showed significant associations only at the network-level. It might be related to this fact that the neurobiological effects of tDCS is less straightforward at the voxels/regions/clusters level [34] and emphesizes the importance of network-level changes during stimulation. Over the past several years there has been a strong movement toward network-based effects of brain stimulation technologies [35] and our results suggest that tDCS effects may tend to spread across the networks not a restricted local brain region. (2) Furthermore, at the network level, we found significant correlations in two different directions; negative correltion during resting-state fMRI inside DMN, and positive correlation during task-based fMRI inside ECN. Based on previous neuroimaging findings DMN exibits a higher activity at resting-state while ECN is commonly active during the goal-oriented tasks [36] we also found significant correltion in DMN during resting-state and in ECN while people were exposed to drug vs neutral cues. Furthermore, the opposite direction of correlations in ECN and DMN could be related to the direction of the electric field in these networks [37]. Furthermore, it could be related to the counterbalance interaction between DMN and ECN activity/connectivity such that reduced functional activity in DMN is associated with increased activation in the task-positive regions such as ECN [38, 39].

#### Challenges that may affect dose-response results

Several challenges exist in integrating EF with fMRI data. In the following section, we primarily consider potential challenges in combining EF distribution patterns measured with computational head models with a functional maps obtained from fMRI data analysis.

##### 1- Integrating a static model with a dynamic map

The actual relationship between tES-induced EFs and functional brain state is not completely clear. Changes in functional activity depend on how a targeted brain region responds to the injected current and how other brain areas interact with the stimulation target. Distant brain regions interact with each other through brain networks. With respect to the activity-selective mechanism of action in tES, activation in one brain region may affect sensitivity to injected current in other functionally connected brain regions and because of the complicated mosaic of interactions between targeted and non-targeted brain regions, it would be difficult to find a linear relationship between EFs and changes in BOLD signals especially in a predefined cortical target such as the cortex underlying the active electrode.

##### 2- Interpretation of the results

Interpretation of the integration results between EF and fMRI would be complicated and controversial since the brain regions are structurally and functionally interconnected. Interpretation of the positive or negative correlation between EFs and changes in the BOLD signal would be complex. For example, if one finds a negative correlation between EFs and changes in fMRI data for a group of subjects, it’s not clear why individuals with higher induced field strengths showed a reduction in connectivity in targeted brain regions during stimulation while those with low induced field strengths showed increased connectivity in the same regions.

##### 3- Selecting an appropriate measure

Different available metrics can be extracted from EF head models or fMRI data. For example, both tangential and normal components of the EFs can be calculated over the cortex. Each provides different information about EF distribution patterns; the tangential EF component is informative of the EF strength while the EF component normal to the cortical surface is informative of EF direction. Furthermore, each component requires a decision on if to use maximum, averaged, median, or some other measures, which can be integrated with fMRI. Similarly, finding a reliable and accurate fMRI measure would be challenging. Any functional map obtained from different resting-state (e.g., functional activity or connectivity during rest) or task-based fMRI analysis (based on different approaches such as model-based or data-driven) can be used for the integration. Each measure leads to different integration results and interpretation, and with respect to the research question, selecting appropriate indices would be challenging.

##### 4- Accuracy of the modeling

Another challenge we face in integrating CHMs with fMRI is the importance of head model precision. Inappropriate modeling of the head or brain leads to an incorrect distribution of the EF in the brain that may alter the results of integration. Any individualized CHM is limited by the precision and accuracy of the tissue segmentation and the assigned conductivity [40]. For example, previous findings suggest that accurate segmentation (ranges from brain tissue to non-brain tissue) would lead to a more accurate personalized brain stimulation [41-44]. In this context, advanced methods were proposed recently to increase the accuracy of CHMs by considering more precise segmentation methods [45-47].

Furthermore, CHMs are commonly generated based on previously established isotropic conductivities for tissues. However, previous studies suggest that inter-individual variability in tissue conductivities [48], anisotropic conductivities (especially for skull and white matter based on using diffusion tensor imaging (DTI)) [49], and nonlinear dynamic changes of tissue-electrode conductivities [50] may affect the accuracy of the CHMs. Additionally, multi-scale models that combine MRI-derived CHMs with multi-compartmental models of cortical neurons and constructed realistic fiber models using DTI have been developed to predict stimulation effects more precisely [51-53]. Taken together, creating CHMs as accurately as possible may be beneficial to find more accurate relationships between EFs and fMRI data.

##### 5- Integration space

Combination can be performed in two different levels as follows: (1) subject space, where two maps are based on the subject space coordination and each CHM combined with own individual fMRI map and this integration is repeated for each participant, and (2) standard space, where CHMs and functional maps are transformed to a standard space (e.g., MNI, Talairach, fsaverage). Although group-level integration provides a comparative analysis of the results across the population, the normalization to standard space could change the EF as it warps each head to the standard head. Instead of generating CHMs for each individual and transformation to standard space that could be a time-consuming procedure, another alternative way in integrating CHMs with fMRI studies is using a standard model such as an arbitrary individual model proposed by Holmes et al. called Collin27 based on 27 MRI scans of a single subject or a standard brain proposed by Huang et al [54, 55]. Although these models reduce susceptibility to single head anatomy compared to using a single subject head model and decrease computational time compared to generating CHMs for each participant, the accuracy of integrating template models with a group-level functional map is lower than the use of personalized models based on individual scans, which will be certainly necessary in specific cases with brain damages such as stroke.

#### Limitations and future directions

Although the current and previous dose-response relatrionship studies for determining the associations between EFs and brain functions are promising, much work remains to be done. Integrating CHMs with fMRI is still new in tES studies, and previous works reported novel findings in this combination. Therefore, replication is needed to draw a general conclusion about the relationship between EF distribution patterns and changes in BOLD signals.

Additionally, thanks to the advancement of computational resources and machine learning techniques like deep learning, it is becoming increasingly easier to deal with more complex nonlinear/higher-order relationships between EFs and functional activity/connectivity at the subject or group level. For example, CHMs and fMRI data can be fed into a deep learning model for classification/prediction purposes. With regard to sufficient data for training, the machine learning methods can be used to classify the participants into two groups; responders and non-responders based on EFs at targeted on non-targeted brain regions and the initial state of the brain. This approach may also help to identify multi-modal biomarkers based on the predictive role of baseline brain activation and individualized EFs.

Closed-loop tES-fMRI studies, where fMRI and tES are performed simultaneously, raise the interesting possibility of real-time integration of personalized ongoing brain function with EFs. This approach can help to optimize stimulation parameters based on how EFs interact with neuronal activity. In the context of integrating CHMs with concurrent fMRI, a functional link can be defined between stimulation parameters and changes in the functional state of the brain. However, online processing of CHMs and fMRI data is a time-consuming procedure. A high-speed algorithm and powerful computational tools are needed to implement a closed-loop CHM-fMRI integration in future studies.

Finally, other types of modalities, including EEG, DTI, or behavioral data, can be integrated with CHMs and fMRI in future studies. Integrating multi-sources of information allows us to organize a hybrid activation pattern to find putative trait-state relationships that might be much more informative than using a single source.

## Conclusion

The proposed multi-level analytic pipeline provides a methodological framework to analyze tDCS effects in terms of dose-response relationships at four different levels to directly link the electric field (received dose) variability to the variability of the neural response to tDCS (response). The exploratory results suggest that network-based analysis might be a better approach to provide novel insights into the dependency of the neuromodulatory effects of tDCS on the brain’s regional current dose in each individual. Dose-response integration can be informative for dose optimization/customization or predictive/treatment-response biomarker extraction in future brain stimulation studies.

## Data Availability

All data produced in the present study are available upon reasonable request to the authors.

## References

1. Mosayebi-Samani, M., et al., The impact of individual electrical fields and anatomical factors on the neurophysiological outcomes of tDCS: A TMS-MEP and MRI study. Brain Stimulation, 2021. 14(2): p. 316–326.

2. Nitsche, M.A. and W. Paulus, Excitability changes induced in the human motor cortex by weak transcranial direct current stimulation. The Journal of physiology, 2000. 527(3): p. 633–639.

3. Regner, G.G., et al., Preclinical to clinical translation of studies of transcranial direct-current stimulation in the treatment of epilepsy: a systematic review. Frontiers in neuroscience, 2018. 12: p. 189.

4. Palm, U., et al., tDCS for the treatment of depression: a comprehensive review. European archives of psychiatry and clinical neuroscience, 2016. 266(8): p. 681–694.

5. Kim, J.-H., et al., Inconsistent outcomes of transcranial direct current stimulation may originate from anatomical differences among individuals: electric field simulation using individual MRI data. Neuroscience letters, 2014. 564: p. 6–10.

6. Lauro, L.J.R., et al., TDCS increases cortical excitability: direct evidence from TMS–EEG. Cortex, 2014. 58: p. 99–111.

7. Chan, M.M. and Y.M. Han, The effect of transcranial direct current stimulation in changing resting-state functional connectivity in patients with neurological disorders: a systematic review. Journal of central nervous system disease, 2020. 12: p. 1179573520976832.

8. Esmaeilpour, Z., et al., Methodology for tDCS integration with fMRI. Human brain mapping, 2020. 41(7): p. 1950–1967.

9. Li, L.M., K. Uehara, and T. Hanakawa, The contribution of interindividual factors to variability of response in transcranial direct current stimulation studies. Frontiers in cellular neuroscience, 2015. 9: p. 181.

10. Hill, A.T., P.B. Fitzgerald, and K.E. Hoy, Effects of anodal transcranial direct current stimulation on working memory: a systematic review and meta-analysis of findings from healthy and neuropsychiatric populations. Brain stimulation, 2016. 9(2): p. 197–208.

11. Horvath, J.C., J.D. Forte, and O. Carter, Quantitative review finds no evidence of cognitive effects in healthy populations from single-session transcranial direct current stimulation (tDCS). Brain stimulation, 2015. 8(3): p. 535–550.

12. Wiethoff, S., M. Hamada, and J.C. Rothwell, Variability in response to transcranial direct current stimulation of the motor cortex. Brain stimulation, 2014. 7(3): p. 468–475.

13. Laakso, I., et al., Can electric fields explain inter-individual variability in transcranial direct current stimulation of the motor cortex? Scientific reports, 2019. 9(1): p. 1–10.

14. Laakso, I., et al., Inter-subject variability in electric fields of motor cortical tDCS. Brain stimulation, 2015. 8(5): p. 906–913.

15. Gomez-Tames, J., et al., Group-level and functional-region analysis of electric-field shape during cerebellar transcranial direct current stimulation with different electrode montages. Journal of neural engineering, 2019. 16(3): p. 036001.

16. Li, L.M., et al., Brain state and polarity dependent modulation of brain networks by transcranial direct current stimulation. Human brain mapping, 2019. 40(3): p. 904–915.

17. Datta, A., et al., Gyri-precise head model of transcranial direct current stimulation: improved spatial focality using a ring electrode versus conventional rectangular pad. Brain stimulation, 2009. 2(4): p. 201-207. e1.

18. Huang, Y., et al., Measurements and models of electric fields in the in vivo human brain during transcranial electric stimulation. Elife, 2017. 6: p. e18834.

19. Opitz, A., et al., Physiological observations validate finite element models for estimating subject-specific electric field distributions induced by transcranial magnetic stimulation of the human motor cortex. Neuroimage, 2013. 81: p. 253–264.

20. Edwards, D., et al., Physiological and modeling evidence for focal transcranial electrical brain stimulation in humans: a basis for high-definition tDCS. Neuroimage, 2013. 74: p. 266–275.

21. Jog, M.V., et al., In-vivo imaging of magnetic fields induced by transcranial direct current stimulation (tDCS) in human brain using MRI. Scientific reports, 2016. 6(1): p. 1–10.

22. Kasten, F.H., et al., Integrating electric field modelling and neuroimaging to explain variability of low intensity tES effects. BioRxiv, 2019: p. 581207.

23. Lang, S., et al., Preoperative transcranial direct current stimulation in glioma patients: a proof of concept pilot study. Frontiers in neurology, 2020. 11.

24. Antonenko, D., et al., Towards precise brain stimulation: Is electric field simulation related to neuromodulation? Brain stimulation, 2019. 12(5): p. 1159–1168.

25. Esmaeilpour, Z., et al., Methodology for tDCS integration with fMRI. Human Brain Mapping, 2019.

26. Jamil, A., et al., Current intensity-and polarity-specific online and aftereffects of transcranial direct current stimulation: An fMRI study. Human brain mapping, 2020. 41(6): p. 1644–1666.

27. Ekhtiari, H., et al., Transcranial direct current stimulation to modulate fMRI drug cue reactivity in methamphetamine users: a randomized clinical trial. medRxiv, 2021.

28. Fan, L., et al., The human brainnetome atlas: a new brain atlas based on connectional architecture. Cerebral cortex, 2016. 26(8): p. 3508–3526.

29. Halko, M., et al., Neuroplastic changes following rehabilitative training correlate with regional electrical field induced with tDCS. Neuroimage, 2011. 57(3): p. 885–891.

30. Heller, R., et al., Cluster-based analysis of FMRI data. NeuroImage, 2006. 33(2): p. 599–608.

31. Bassett, D.S. and O. Sporns, Network neuroscience. Nature neuroscience, 2017. 20(3): p. 353–364.

32. Soleimani, G., et al., Group and individual level variations between symmetric and asymmetric DLPFC montages for tDCS over large scale brain network nodes. Scientific reports, 2021. 11(1): p. 1–13.

33. Indahlastari, A., et al., High fidelity finite element models to predict changes in functional connectivity of the working memory networks in older adults. Brain Stimulation: Basic, Translational, and Clinical Research in Neuromodulation, 2021. 14(6): p. 1652.

34. Leaver, A.M., et al., Modulation of brain networks during MR-compatible transcranial direct current stimulation. Neuroimage, 2022. 250: p. 118874.

35. Beynel, L., J.P. Powers, and L.G. Appelbaum, Effects of repetitive transcranial magnetic stimulation on resting-state connectivity: A systematic review. Neuroimage, 2020. 211: p. 116596.

36. Park, H.-J. and K. Friston, Structural and functional brain networks: from connections to cognition. Science, 2013. 342(6158): p. 1238411.

37. Soleimani, G., et al., DLPFC stimulation alters large-scale brain networks connectivity during a drug cue reactivity task: A tDCS-fMRI study. Frontiers in systems neuroscience, 2022. 16.

38. Wang, R.W., et al., Posterior cingulate cortex can be a regulatory modulator of the default mode network in task-negative state. Scientific Reports, 2019. 9(1): p. 1–12.

39. Fox, M.D., et al., The human brain is intrinsically organized into dynamic, anticorrelated functional networks. Proceedings of the National Academy of Sciences, 2005. 102(27): p. 9673–9678.

40. Bikson, M. and A. Datta, Guidelines for precise and accurate computational models of tDCS. Brain stimulation, 2012. 5(3): p. 430.

41. Lee, E.G., et al., Impact of non-brain anatomy and coil orientation on inter-and intra-subject variability in TMS at midline. Clinical Neurophysiology, 2018. 129(9): p. 1873–1883.

42. Janssen, A., et al., The influence of sulcus width on simulated electric fields induced by transcranial magnetic stimulation. Physics in Medicine & Biology, 2013. 58(14): p. 4881.

43. Opitz, A., et al., How the brain tissue shapes the electric field induced by transcranial magnetic stimulation. Neuroimage, 2011. 58(3): p. 849–859.

44. Thielscher, A., A. Opitz, and M. Windhoff, Impact of the gyral geometry on the electric field induced by transcranial magnetic stimulation. Neuroimage, 2011. 54(1): p. 234–243.

45. Jiang, J., et al., Enhanced tES and tDCS computational models by meninges emulation. Journal of neural engineering, 2020. 17(1): p. 016027.

46. Rashed, E.A., J. Gomez-Tames, and A. Hirata, Influence of segmentation accuracy in structural MR head scans on electric field computation for TMS and tES. Physics in Medicine & Biology, 2021. 66(6): p. 064002.

47. Nielsen, J.D., et al., Automatic skull segmentation from MR images for realistic volume conductor models of the head: Assessment of the state-of-the-art. Neuroimage, 2018. 174: p. 587–598.

48. Schmidt, C., et al., Impact of uncertain head tissue conductivity in the optimization of transcranial direct current stimulation for an auditory target. Journal of neural engineering, 2015. 12(4): p. 046028.

49. Suh, H.S., W.H. Lee, and T.-S. Kim, Influence of anisotropic conductivity in the skull and white matter on transcranial direct current stimulation via an anatomically realistic finite element head model. Physics in Medicine & Biology, 2012. 57(21): p. 6961.

50. Bikson, M., et al., Transcranial electrical stimulation nomenclature. Brain stimulation, 2019. 12(6): p. 1349–1366.

51. Seo, H. and S.C. Jun, Multi-scale computational models for electrical brain stimulation. Frontiers in human neuroscience, 2017. 11: p. 515.

52. Seo, H., et al., A multi-scale computational model of the effects of TMS on motor cortex. F1000Res. 5, 1945. 2017.

53. Shirinpour, S., et al., Multi-scale Modeling Toolbox for Single Neuron and Subcellular Activity under (repetitive) Transcranial Magnetic Stimulation. BioRxiv, 2020.

54. Holmes, C.J., et al., Enhancement of MR images using registration for signal averaging. Journal of computer assisted tomography, 1998. 22(2): p. 324–333.

55. Huang, Y., L.C. Parra, and S. Haufe, The New York Head—A precise standardized volume conductor model for EEG source localization and tES targeting. NeuroImage, 2016. 140: p. 150–162.

